# OphthaBERT: Automated Glaucoma Diagnosis from Clinical Notes

**DOI:** 10.1101/2025.06.08.25329151

**Authors:** Rishi Shah, Mousa Moradi, Sedigheh Eslami, Asahi Fujita, Kanza Aziz, Niloufar Bineshfar, Tobias Elze, Mohammad Eslami, Saber Kazeminasab, Daniel Liebman, Saeid Rasouli, Daniel Vu, Mengyu Wang, Jithin Yohannan, Nazlee Zebardast

## Abstract

Glaucoma is a leading cause of irreversible blindness worldwide, with early intervention often being crucial. Research into the underpinnings of glaucoma often relies on electronic health records (EHRs) to identify patients with glaucoma and their subtypes. However, current methods for identifying glaucoma patients from EHRs are often inaccurate or infeasible at scale, relying on International Classification of Diseases (ICD) codes or manual chart reviews. To address this limitation, we introduce (1) OphthaBERT, a powerful general clinical ophthalmology language model trained on over 2 million diverse clinical notes, and (2) a fine-tuned variant of OphthaBERT that automatically extracts binary and subtype glaucoma diagnoses from clinical notes. The base OphthaBERT model is a robust encoder, outperforming state-of-the-art clinical encoders in masked token prediction on out-of-distribution ophthalmology clinical notes and binary glaucoma classification with limited data. We report significant binary classification performance improvements in low-data regimes (p < 0.001, Bonferroni corrected). OphthaBERT is also able to achieve superior classification performance for both binary and subtype diagnosis, outperforming even fine-tuned large decoder-only language models at a fraction of the computational cost. We demonstrate a 0.23-point increase in macro-F1 for subtype diagnosis over ICD codes and strong binary classification performance when externally validated at Wilmer Eye Institute. OphthaBERT provides an interpretable, equitable framework for general ophthalmology language modeling and automated glaucoma diagnosis.

## 1 Introduction

Electronic health records (EHRs) contain a wealth of patient health information. While patient health information can be encoded in structural features such as lab results or International Classification of Diseases (ICD) codes, it is estimated that 80% of clinical information is held in unstructured formats such as clinical notes [1]. Data in clinical notes captures the physician’s interpretation of patient information and is often essential for accurately interpreting patient data [2].

Manually extracting information from clinical notes is time-consuming and error-prone, motivating the development of natural language processing (NLP) models to automate this process. Recent machine learning advances, particularly through the development of the transformer family of models, have allowed for a new standard of automated language processing and understanding [3]. Language models optimized for medical domains, such as BioClinicalBERT and Med-PaLM, have proven to capture complex context and relationships in text [4–6]. While these models excel at general clinical reasoning tasks, these generalized pretrained models struggle to capture the nuance required in specialized clinical settings [5].

Ophthalmology is a medical specialty spanning a broad spectrum of eye diseases, surgical techniques, and diagnostic procedures. This complexity results in highly specialized language that is difficult to process [7]. Although showing a promising start, language models such as Med-PaLM and ChatGPT are costly and struggle to perform well in ophthalmic subspecialties due to being pretrained on minimal domain-specific corpora [6, 8]. We aim to develop a lightweight ophthalmology language model to address this gap and accelerate clinical research.

Glaucoma is a leading cause of irreversible blindness worldwide, and early intervention is crucial for improving outcomes [9]. Despite recent advancements in diagnostic and therapeutic techniques, the complex multifactorial nature together with asymptomatic onset and progression of glaucoma renders early detection and personalized treatment a significant challenge. Treatment optimization, the ability to reliably predict disease course, and personalized management are key to preserving visual function. Historically, glaucoma research has relied primarily on ICD codes and manual chart review for case ascertainment. However, the former suffers from inaccuracies, with self-reported cases agreeing with structured EHR documentation only 50% of the time [10]. Reliably extracting glaucoma diagnoses from clinical notes through natural language processing would significantly improve the quantity and quality of labeled patients for future studies, which has the potential to rapidly accelerate glaucoma research.

We introduce OphthaBERT, a domain-specific clinical language model pretrained on more than two million ophthalmology notes that span diverse subspecialties and patient populations. Building on this foundation, we present an end-to-end framework that automatically extracts glaucoma diagnoses and distinguishes disease subtypes directly from free-text clinical documentation, removing a major bottleneck for large-scale studies. Across benchmark tasks OphthaBERT matches, and surpasses, the performance of other language models, while remaining interpretable and requiring only a fraction of the computational resources.

## 2Results

### 2.1 Sample selection

We designed our base model, OphthaBERT, as a flexible pretrained encoder language model that can be utilized to produce domain-aware embeddings for ophthalmology tasks. As a result, OphthaBERT can be fine-tuned for specific tasks or be used to generate pretrained embeddings for downstream models (Figure 1). Our glaucoma model is then built on top of the base model, with a binary classification head for glaucoma diagnosis and a multiclass classification head for subtype differentiation.

**Figure 1:**
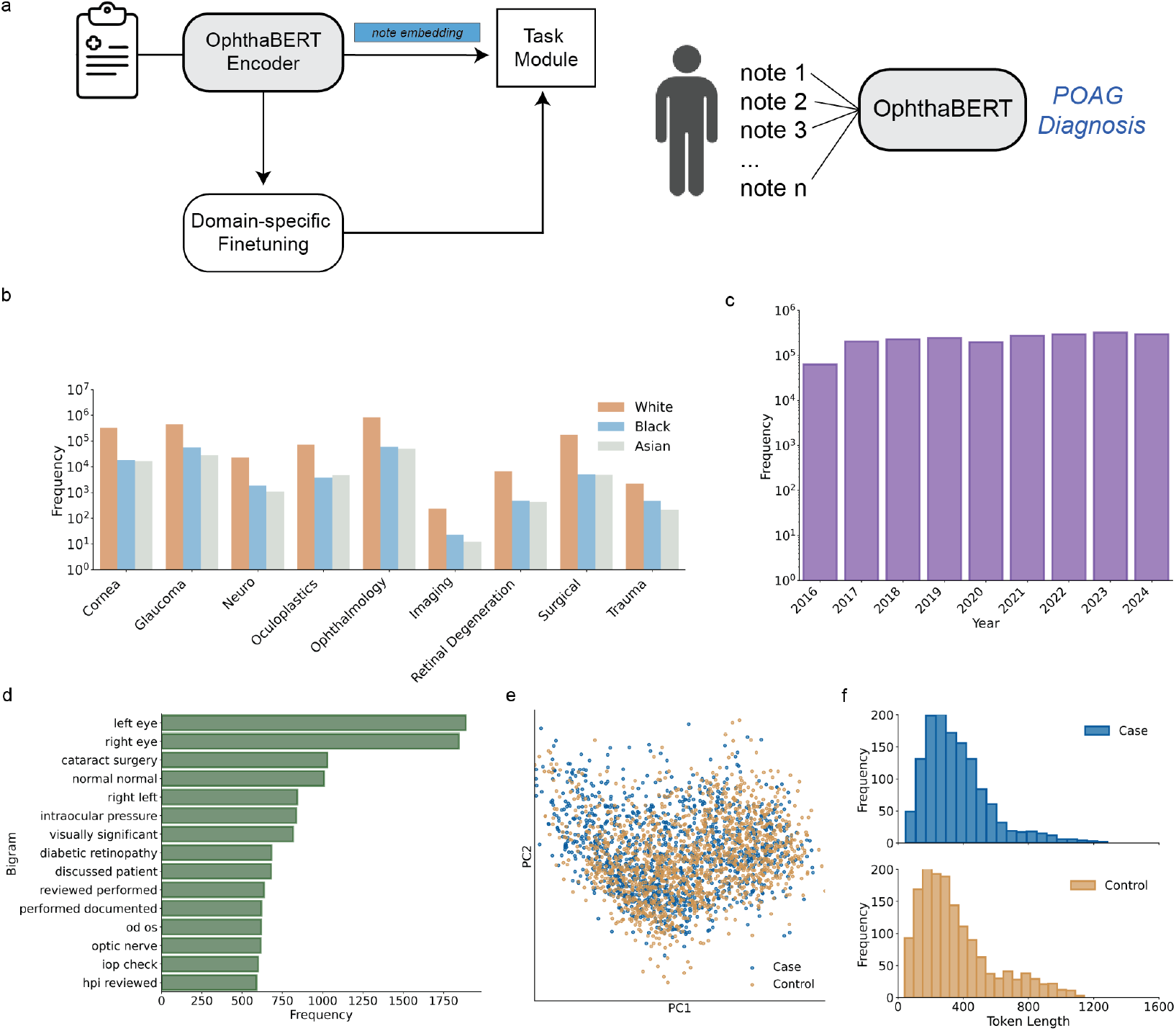
Model architecture and pretraining sample statistics. **(a)** General flowchart for utilizing OphthaBERT in ophthalmology language tasks. OphthaBERT can either be fine-tuned, or the pretrained text embeddings produced by the model can be fed directly into task-specific models for efficient domain-aware encoding. In this work, our task module is predicting glaucoma diagnoses from unstructured clinical notes. **(b)** Distribution of notes from Massachusetts Eye and Ear (MEE) Infirmary by race and subspecialty clinic for masked pretraining of OphthaBERT. **(c)** Distribution of pretraining notes by contact date. **(d)** Frequencies of the top 15 bigrams in labeled notes utilized for downstream glaucoma identification. **(e)** Visualization of the principal components of the embeddings of the [CLS] token for labeled case and control notes before pretraining and fine-tuning OphthaBERT. **(f)** Distribution of note lengths of case and control notes for supervised tuning.

OphthaBERT was pretrained on 2,129,171 de-identified ‘progress’ notes from 327,814 unique patients seen at Massachusetts Eye and Ear (MEE) Infirmary between January 2016 and December 2024 (Figure 1a,b). Our glaucoma model was trained and validated using a distinct subset of 3,003 notes from 1,013 patients annotated by clinicians with both subtype and binary diagnosis labels. Further details of clinical grading are available in the methods section and Supplementary Table 1.

**Table 1:**
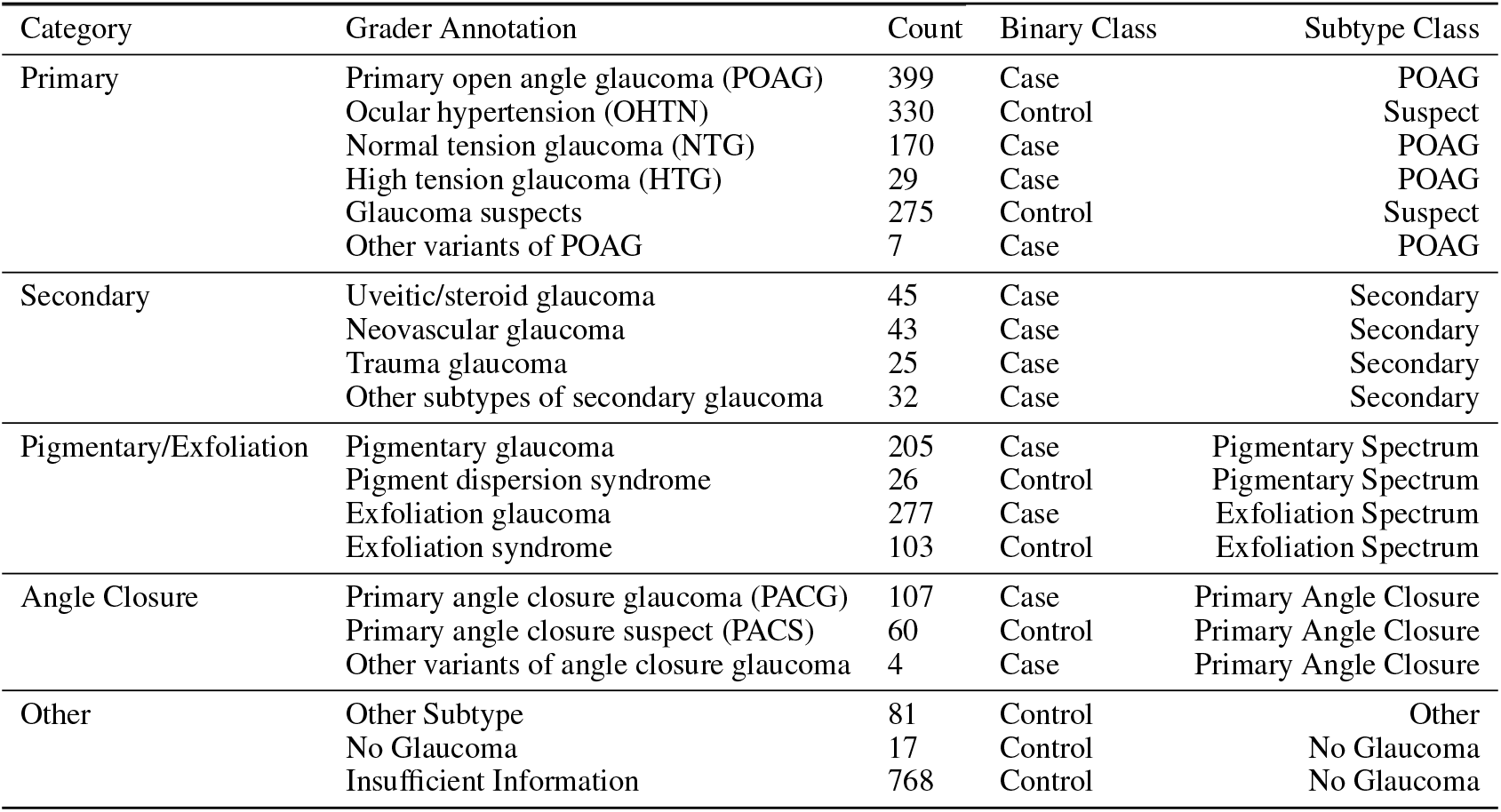
Clinician Labels for Glaucoma Diagnoses and Subtypes.

The most frequent bigrams seen in the notes were ‘left eye’ and ‘right eye’, and a variety of conditions were represented in the notes such as cataracts and diabetic retinopathy (Figure 1d). Notes given binary ‘case’ and ‘control’ labels are structurally similar, indicating that separation between the classes is not trivial. Figure 1e shows substantial overlap between ‘case’ and ‘control’ note embeddings before pretraining and training. Additionally, ‘case’ and ‘control’ note lengths follow similar distributions (Figure 1f).

### 2.2 Masked language modeling pretraining improves ophthalmology-specific representations

Through masked language modeling pretraining, we aimed for our base model, OphthaBERT, to learn ophthalmology-specific representations by predicting masked tokens in clinical notes. Pretraining details are available in the Methods section. We assessed the impact of pretraining through minimalistic downstream fine-tuning for binary glaucoma classification and masked token prediction on an out-of-distribution dataset.

We evaluate the impact of pretraining by adapting OphthaBERT and BioClinicalBERT for binary classification with a simple binary classification head. Although we present a more robust classifier in later results, this modification was designed to be minimal and identical, so that performance differences could be attributed to the choice of the base model after fine-tuning rather than the design of the fine-tuning architecture. We utilize McNemar’s test to evaluate the difference in binary classification performance between OphthaBERT and BioClinicalBERT at various fractions of training data (Figure 2a). OphthaBERT shows substantial improvements over the base model with limited training data. At the 30% and 50% fractions, OphthaBERT’s predictions produce notable increases in macro-F1 score, differing significantly from the base model before pretraining (p < 0.001). The performance of OphthaBERT is also greater at all thresholds, although insignificantly at the lowest and highest thresholds.

**Figure 2:**
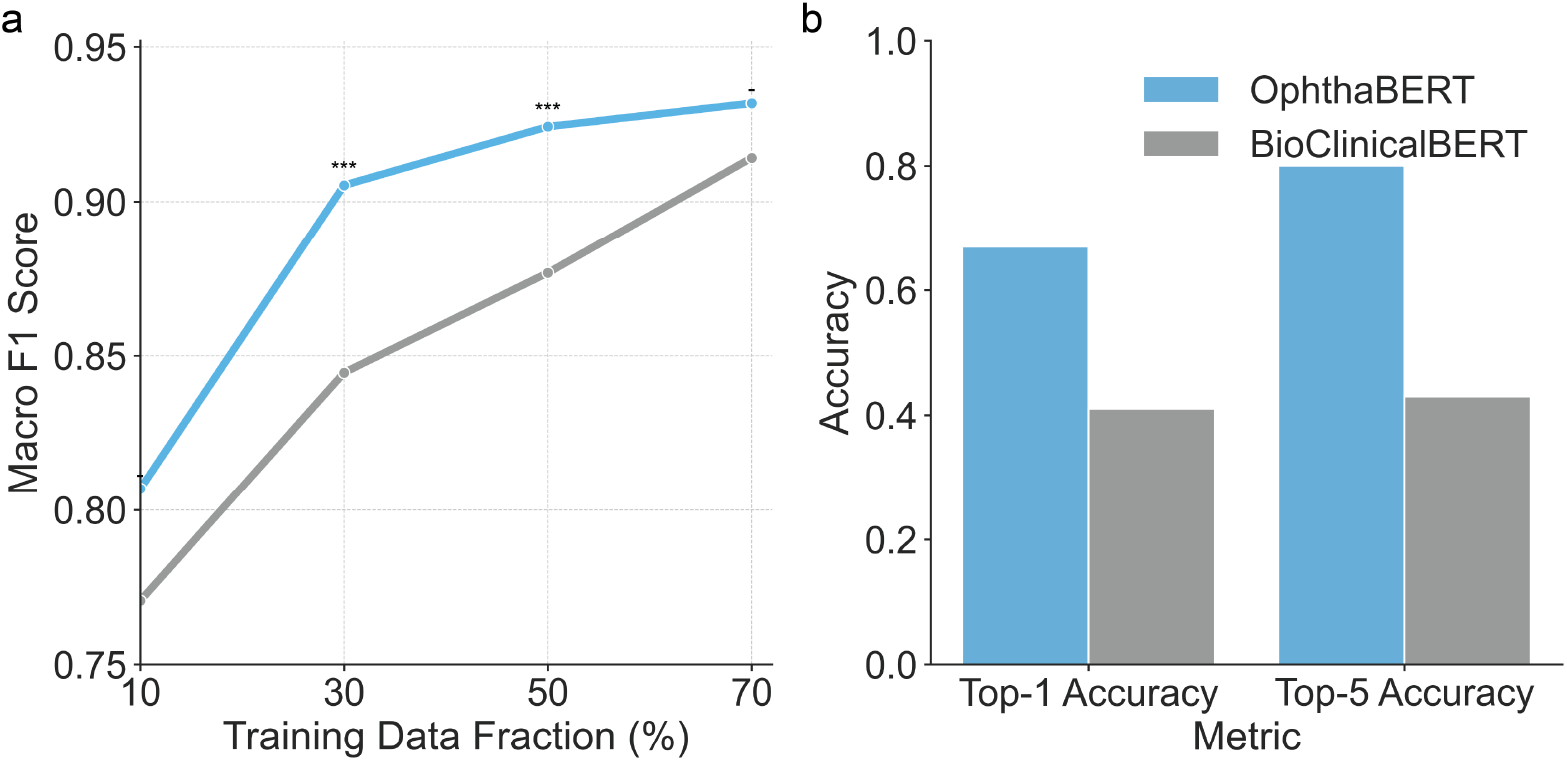
Pretraining improves ophthalmology task performance. **(a)** Binary glaucoma classification macro averaged F1 score across different fractions of labeled data used in training. We attach a simple classification head to the base OphthaBERT model to directly compare the impact of pretraining over specific architecture. Significance between models was evaluated through paired McNemar’s Test, and p-values were corrected by Bonferroni correction to account for multiple hypothesis testing. The thresholds for significance are: n.s. (-), p < 0.05 (*), p < 0.01 (**), p < 0.001 (***) **(b)** Top-1 and top-5 accuracy of OphthaBERT and BioClinicalBERT for masked token prediction on an out-of-distribution dataset. Top-1 accuracy is defined as the model’s accuracy in finding the correct token with one guess. Top-5 accuracy is defined as the accuracy of the model to find the correct token among the top 5 predicted tokens.

We also assess the impact of pretraining through masked token prediction on an out-of-distribution dataset of clinical ophthalmology notes. The dataset used for benchmarking comes from an anonymized set of 480 clinical notes. Patients in this dataset were seen for glaucoma by a comprehensive or glaucoma ophthalmologist from January 1, 2019, to August 31, 2020, at Oregon Health & Science University [11]. OphthaBERT outperforms BioClinicalBERT after pretraining, with an absolute 26% top-1 accuracy increase and a 37% top-5 accuracy increase for predicting masked tokens (Figure 2b).

### 2.3 Glaucoma binary and subtype classification

#### 2.3.1 Equitable glaucoma diagnosis and subtype identification

In this work, we build on the base OphthaBERT model by appending two parallel classification heads: a binary head that discriminates between glaucoma (‘case’) and non-glaucoma (‘control’) notes, and a subtype head that assigns one of seven clinically relevant glaucoma subtypes (non-glaucoma, primary open angle glaucoma (POAG), glaucoma suspect, primary angle closure, exfoliation spectrum, pigmentary spectrum, secondary glaucoma). Architectural details for this model are available in the Methods section. OphthaBERT produces a set of 2 labels for each note, a binary glaucoma diagnosis and a subtype diagnosis. We then utilize a priority-based aggregation system to give each patient a set of 2 diagnoses encompassing all their notes. Model performance is reported through 5-fold cross-validation, splitting on patient ID to prevent data leakage (Figure 3). More details for evaluation by cross-validation are available in the Methods section.

**Figure 3:**
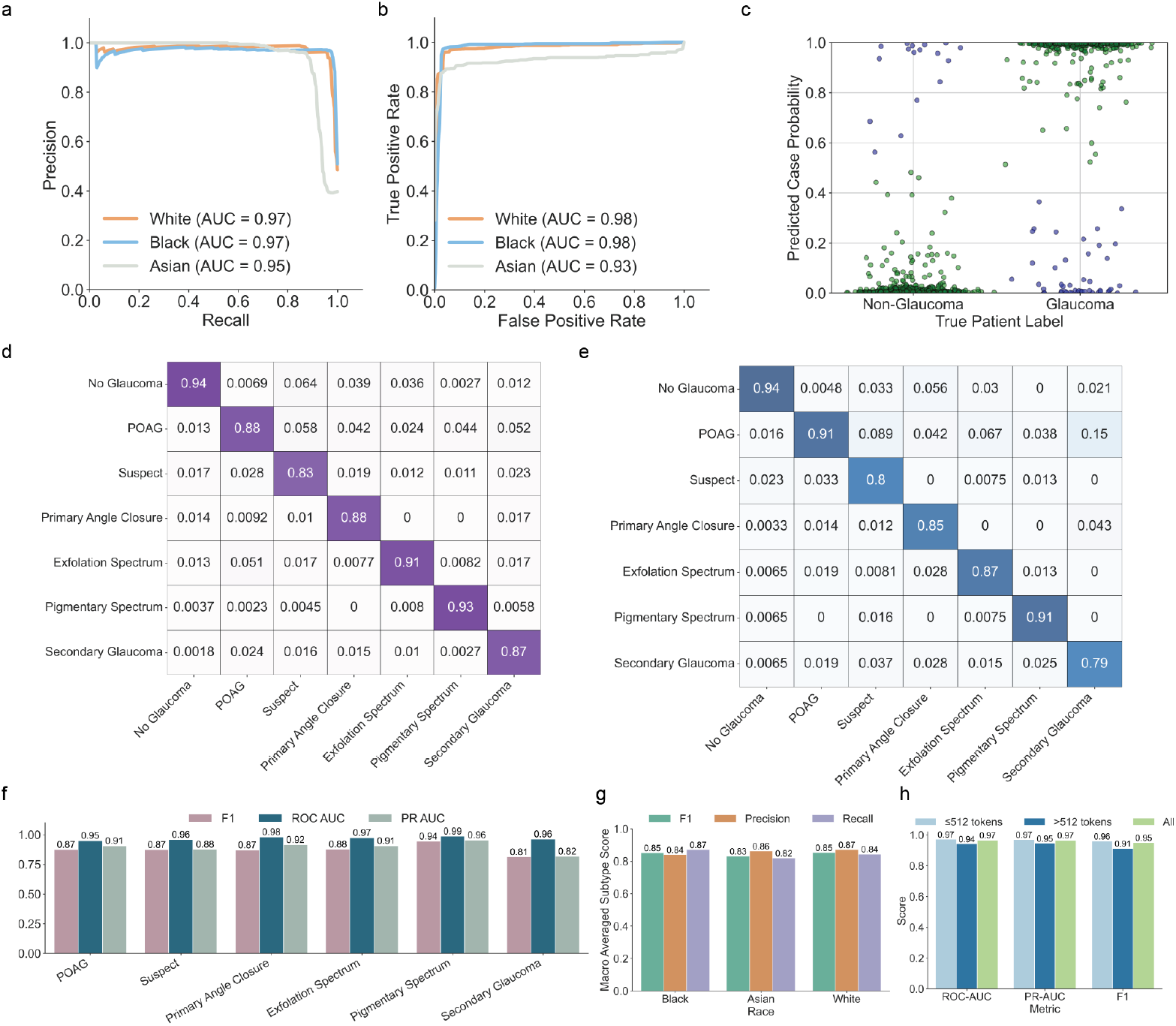
Binary and subtype glaucoma diagnosis performance. (a) Precision-recall curves for each racial group for note-level binary classification. (b) ROC-AUC curves for each racial group for note-level binary classification. (c) Predicted probabilities for glaucoma cases and controls when aggregating note-level labels to produce patient labels. Correct predictions (green) and incorrect predictions (blue) are spread with Gaussian noise along the x-axis for greater clarity. (d) Note prediction-normalized confusion matrix for subtype classification. Predicted classes are reported on the x-axis and true classes are reported on the y-axis. (e) Patient prediction-normalized confusion matrix for subtype classification. Predicted classes are reported on the x-axis and true classes are reported on the y-axis. (f) Note subtype classification performance for each subtype when evaluated against all other classes. (g) Macro-averaged note classification performance for each racial group. (h) Binary note-level classification performance for notes inside and outside of the 512-token BERT context window.

For binary note classification, OphthaBERT’s performance is strong, with ROC-AUC of 0.97 and PR-AUC of 0.97 overall. These results are equitable across all reported racial groups, evidenced by the high, balanced ROC-AUC and PR-AUC values among White, Black, and Asian patients (Figure 3b). Moreover, the model is able to effectively separate ‘case’ and ‘control’ patients, with higher confidence in correct predictions, and lower confidence in incorrect predictions (Figure 3c). Given that BERT has a context window of 512 tokens, we evaluate classification performance of notes both inside and outside the context window to assess the impact of truncation on model performance (Figure 3h). Classification performance remains high for notes inside and outside the context window (ROC-AUC, PR-AUC, F1 > 0.90), suggesting diagnostic information is often present in the first 512 tokens.

We also report confusion matrices for note and patient subtype classification (Figure 3d,e). OphthaBERT is effective at discriminating most subtypes, but struggles most with secondary glaucoma (Figure 3f). This is likely due to secondary glaucoma being defined as a heterogeneous set of conditions with inherently noisy labels. Despite this dip in performance, OphthaBERT still reports ROC-AUC > 0.94 and F1 and PR-AUC values above 0.80 for all subtypes. Importantly, these results are equitable across racial groups. Averaging across all subtypes, the subtype classifier is equitable, achieving macro-averaged precision, recall, and F1 scores all above 0.85 for each racial group (Figure 3g).

#### 2.3.2 Benchmarking and interpretability

We benchmark OphthaBERT’s performance with other models (Figure 4). All models were fine-tuned and evaluated on identical splits. More details are available in the methods section.

**Figure 4:**
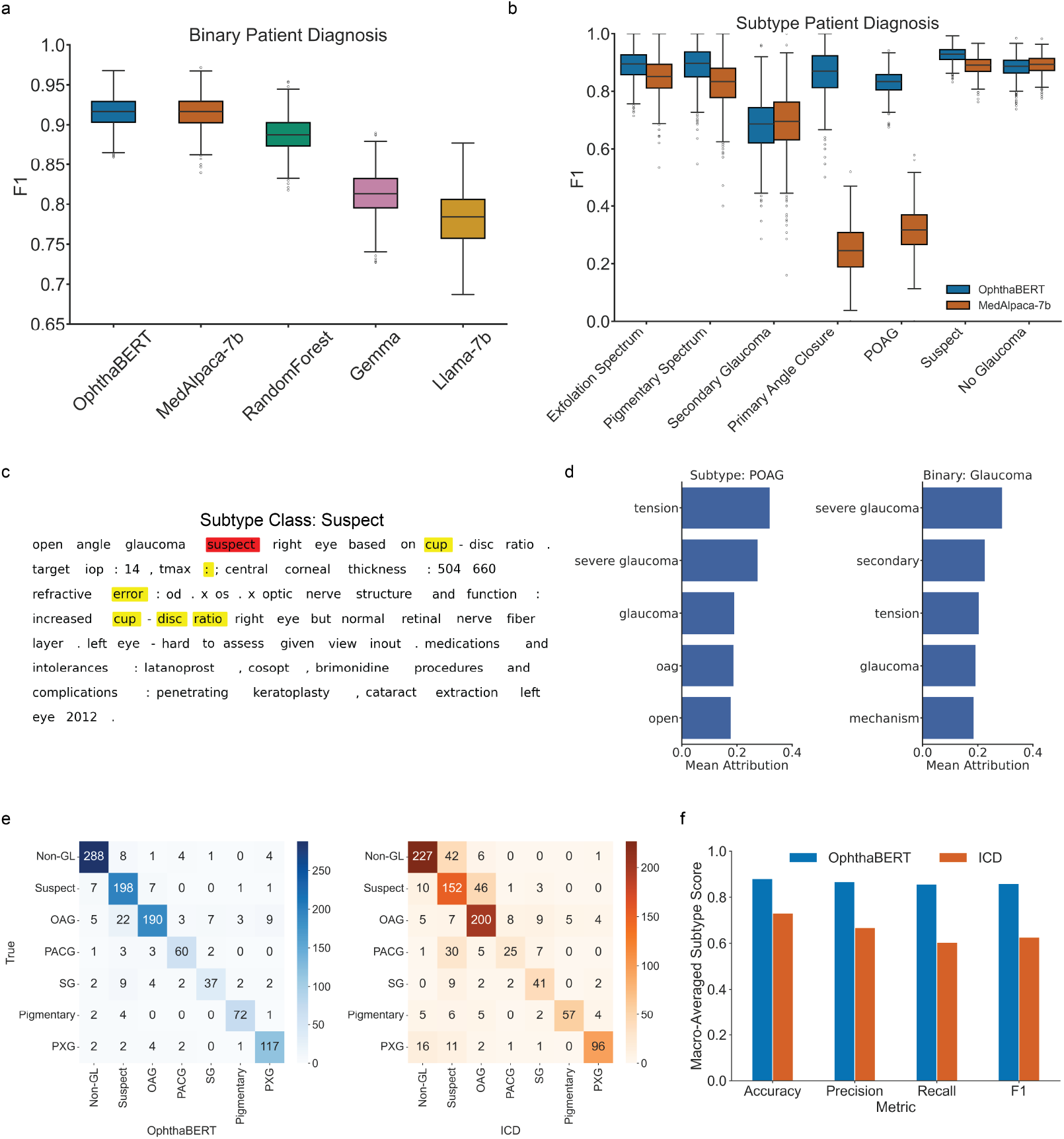
Benchmarking and Interpretability. **(a)** Bootstrapped patient-level binary glaucoma classification performance benchmarked on identical splits. **(b)** Bootstrapped patient-level subtype glaucoma classification performance benchmarked on identical splits. **(c)** Example attributions for subtype classification of a ‘suspect’ note utilizing integrated gradients. **(d)** Words containing tokens with the highest average attributions for notes of primary open angle glaucoma (POAG) subtype labels and glaucoma binary labels. **(e)** Patient-level subtype glaucoma classification performance benchmarked against ICD codes. **(f)** Comparison of patient-level subtype classification between OphthaBERT and ICD codes averaged over all subtypes.

OphthaBERT is competitive with benchmarked models, including state-of-the-art large language models (LLMs) such as MedAlpaca-7B, Gemma-1.1B, and Llama-2-7B (Figure 4a). This performance gain is magnified in subtype classification, where the strongest binary competitor, MedAlpaca-7B, struggles with primary open angle glaucoma (POAG) and primary angle closure subtypes (Figure 4b). OphthaBERT’s improvements come at a fraction of the computational cost, as OphthaBERT has only 2% of the parameters of MedAlpaca-7B and Llama-2-7B. This allows for faster inference with less memory footprint, making OphthaBERT more accessible and practical at the scale of EHR data.

Since glaucoma diagnosis is often inferred through ICD codes in large EHR datasets, we also benchmark OphthaBERT against ICD codes. Utilizing the same priority labeling system we used to assign note-level diagnoses, each patient was given a single ICD code diagnosis. OphthaBERT’s predictions are more accurate and balanced than ICD codes, with a 0.23-point absolute increase in macro-averaged F1-score (Figure 4e,f).

A key advantage of the BERT architecture is that it allows for fine-grained interpretability of predictions. Importance values can be computed down to the sub-word token for each layer of the model. Through our interpretability analysis, we can show exactly which words in notes contribute to a given prediction for both binary and subtype identification. Details for interpretability analysis are available in the Methods section. Figure 4c shows the attributions of words for subtype detection, with the model paying the most attention to critical words such as ‘suspect’ for its prediction. We average token-level attribution scores across all notes for subtype and binary classification heads and report the most influential words for ‘POAG’ subtype diagnosis and ‘Glaucoma’ binary diagnosis (Figure 4d).

#### 2.3.3 External Validation

We externally validated the binary classification module of OphthaBERT on a set of 70 notes from Johns Hopkins University (JHU) Department of Ophthalmology. This dataset consisted of 35 definitive glaucoma notes, 9 possible glaucoma notes, and 26 non-glaucoma notes.

We examine the model’s performance across different note types, including Overview Notes, Progress Notes, and Assessment and Plan Notes (A&P) concatenated with Overview Notes (Figure 5). We also compare the model’s performance when including and excluding notes given ‘Possible Glaucoma’ labels. For ‘A&P + Overview’ and ‘Progress’ notes, the model performance achieves high performance across various probability thresholds (Figure 5a). Most of the uncertainty in the model’s predictions comes from the presence of notes with ‘possible’ labels, as evidenced by the slight drop in performance when including these notes. However, this performance dip is expected, as ‘Possible Glaucoma’ diagnoses are often highly subjective, with different criteria on what can be treated as a ‘Definitive Glaucoma’ case.

**Figure 5:**
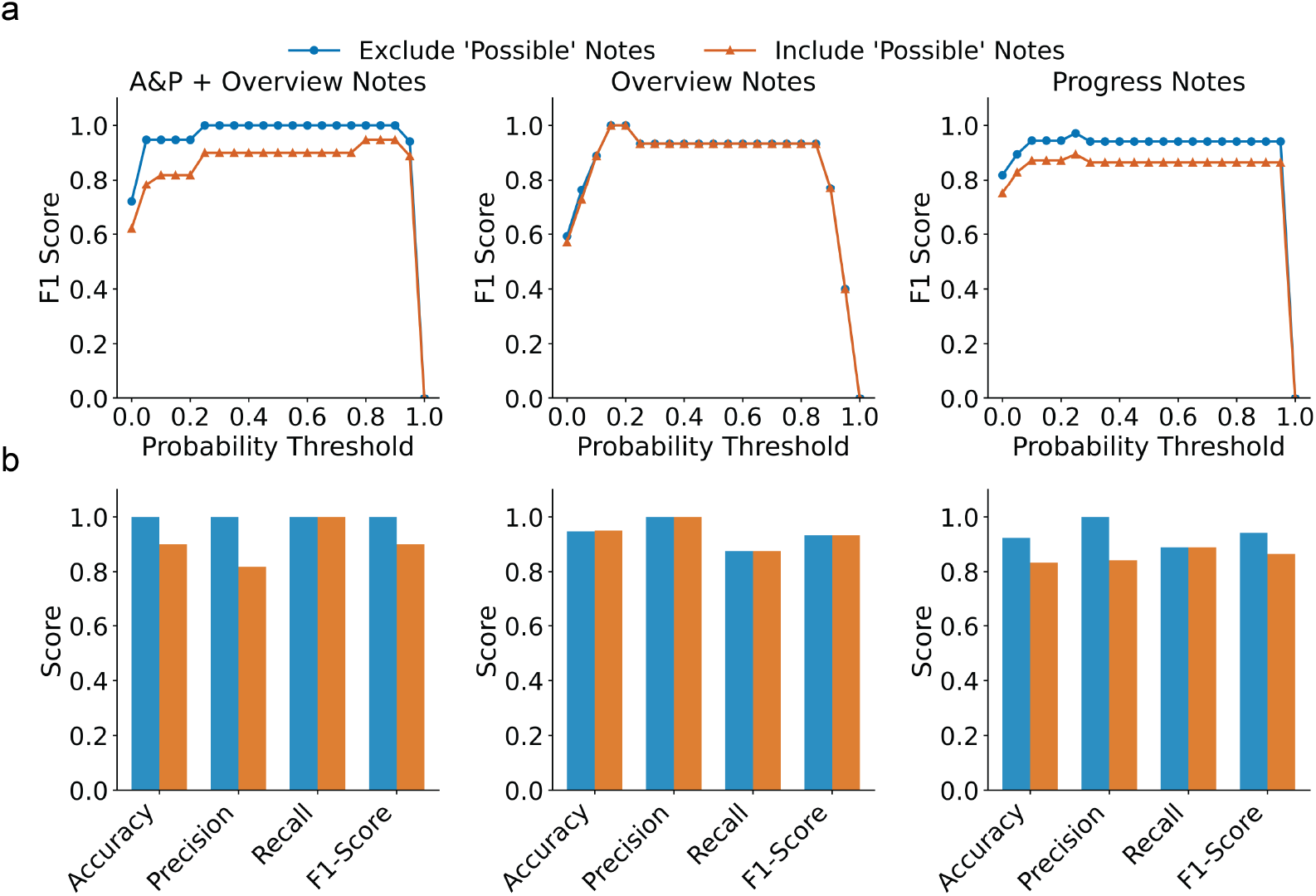
Hopkins External Validation. **(a)** Binary note F1 score at various probability cutoff thresholds for a set of 70 notes from Johns Hopkins University (JHU) Department of Ophthalmology. We examine the performance of the model when including and excluding notes with ‘possible glaucoma’ labels for various note types. We examined performance on assessment and plan notes concatenated with overview notes (A&P + Overview), overview notes, and progress notes. **(b)** Various performance metrics for OphthaBERT’s binary glaucoma diagnosis on the JHU dataset.

We also report various other performance metrics for OphthaBERT’s binary glaucoma diagnosis on the JHU dataset (Figure 5b). The results are consistent with the findings above, showing strong and balanced performance across all note types.

## 3 Discussion

In this study, we developed OphthaBERT, a novel domain-optimized language model for clinical ophthalmology tasks. This work builds on the general clinical language model BioClinicalBERT to learn complex clinical ophthalmology representations. Previous work has trained ophthalmology language models on abstracts, textbooks, and articles (EYE-Llama), but this study presents a novel ophthalmology language model pretrained explicitly for clinical tasks [12]. Other clinical language models are pretrained on biased generic clinical notes, such as MIMIC-III, a dataset of hospital discharge notes with over 80% white patients. Prioritizing racial equity and specialized ophthalmology performance, our model is pretrained on a robust set of notes representing patients from diverse ophthalmology departments and racial groups.

Through OphthaBERT, we introduce a methodology for developing text-based machine learning methods within ophthalmology. As was done in this work, we suggest fine-tuning OphthaBERT for individual downstream use cases with limited labeled data. While existing large language models are resource intensive to fine-tune or prompt for niche tasks, OphthaBERT serves as a lightweight and accessible domain-optimized model. Our results show that OphthaBERT is more effective than existing state-of-the-art competitive models in downstream glaucoma diagnosis with limited data, highlighting its practical utility in low-data regimes.

Studies have used the embeddings produced by BERT, ClinicalBERT, and BioClinicalBERT to feed into specialized or multimodal models [13]. For those tasks, we suggest replacing embeddings produced by other variants of BERT with the pretrained embeddings produced by OphthaBERT.

In addition to the base OphthaBERT model, we present a fine-tuned variant of OphthaBERT for glaucoma diagnosis and subtype detection, which is a substantial advancement in automated glaucoma detection from EHRs. Many existing models for glaucoma detection are dependent on structural data, making generalization to other EHR structures challenging. Existing NLP approaches also primarily only target specific subtypes of glaucoma, such as primary open angle glaucoma [14].

This study provides a strong dual classifier optimized for both identifying cases of glaucoma in EHRs and differentiating between numerous glaucoma subtypes. As our work utilizes an encoder-based architecture, OphthaBERT’s predictions are interpretable and provide granularity at the word level. Existing methods for identifying glaucoma cases and subtypes often utilize inconsistent ICD codes, which are aggregate labels over an entire patient’s history. We show that our model outperforms this baseline while providing greater granularity through note-level predictions. Through integration with large EHR datasets and biobanks, OphthaBERT has the potential to rapidly accelerate glaucoma research through more accurate, efficient, and equitable identification of glaucoma cases and their subtypes.

Although promising, a limitation of this work is the 512-token context window of OphthaBERT. As a result, OphthaBERT is unable to process very long notes effectively. However, this tradeoff comes with increased efficiency and accessibility of the model, with minimal performance drops. In practice, most notes fall within this context window or contain relevant information in the first 512 tokens. Future work should explore further validating OphthaBERT for more advanced and diverse tasks.

## 4 Methods

### 4.1 MEE Corpus Curation

To create the Mass Eye and Ear (MEE) corpus, we selected 2,129,171 de-identified notes from 327,814 unique patients seen at 9 departments at the MEE Ophthalmology practice between January 2016 and December 2024. In order to reduce racial bias, notes were sampled from three separate racial groups: White, Black, and Asian. We also prioritized sampling notes from patients seen at many different sub-departments, including glaucoma, retina, cornea, oculoplastics, and neuro-ophthalmology.

To preserve the rich contextual information leveraged by bidirectional transformers, we applied minimal processing to the text. All notes were fully anonymized by using the HIPAA-compliant, open-source library Philter. Philter uses several rule-based filters to de-identify names. We added additional rules to remove the names of patients and physicians at MEE. This process automated the removal of all Protected Health Information (PHI), such as names, dates, addresses, and any other potentially identifiable information. In addition, we removed non-punctuation special characters, separated sentences into new lines, and converted all characters to lowercase.

### 4.2 Glaucoma Dataset Curation

In the second phase, we sampled a set of 3,003 notes from a diverse set of 1,013 unique patients for clinician grading. Notes were sampled using a stratified random sampling strategy in order to ensure balanced representation across racial groups (White, Black, Asian) and glaucoma subtypes, including primary open angle glaucoma (POAG), primary angle-closure glaucoma (PACG), pigmentary glaucoma (PDG), exfoliation glaucoma (PXG), and other forms of glaucoma. Sampling was balanced through keyword selection. We kept this balance in order to minimize racial bias and ensure sufficient signal for rare subtypes. Sampled notes were then given to a team of clinicians for manual annotation.

Each clinician identified whether each note diagnosed a patient with glaucoma, along with the corresponding subtype for the diagnosis. Specific counts of subtypes and labels are available in Supplementary Material Table 1. We aimed to sample a representative set of conditions seen at a glaucoma clinic, including borderline cases such as ‘glaucoma suspects’ and a wide variety of ocular subtypes (Figure 1d-f). These notes were then binned into more generalizable ‘subtype’ and ‘binary’ classes, which were used as the training classes for our model (Supplementary Table 1).

#### 4.2.1 Clinician Grading of Glaucoma Dataset

A team of 6 clinicians assigned binary and subtype labels to each note based on the presence or absence of a glaucoma diagnosis documented anywhere in the note. In order to assess inter-annotator reliability, each note was assigned to 2 graders. For notes with disagreements in labels, we kept the majority label. The Cohen’s Kappa statistic was calculated to assess the level of agreement between the annotators. We report a Kappa value of 0.85 in our clinicians’ grades.

We consolidated manual labels into two binary categories: cases and controls. Cases were reserved exclusively for glaucoma-diagnosing subtypes: primary open angle glaucoma (POAG), normal tension glaucoma (NTG), high tension glaucoma (HTG), secondary glaucoma, pigmentary glaucoma, exfoliation glaucoma, and angle closure glaucoma. All other cases of notes were treated as ‘controls’ for the purpose of binary labeling. This includes challenging borderline cases such as glaucoma suspects and pigmentary/exfoliation syndromes as well as notes with insufficient information to discern a diagnosis. We allowed for greater granularity in subtype labeling. All subtypes of POAG (NTG, HTG) were grouped as POAG. Patients with ocular hypertension were grouped with glaucoma suspects as these patients presented with elevated risk for glaucoma without having optic nerve damage. All subtypes of secondary glaucoma were grouped into a single label. Pigmentary glaucoma and pigmentary syndrome were grouped together as pigmentary spectrum for subtype classification but were given different binary labels. This same process was repeated for exfoliation spectrum. Graders were also given the option to label a subtype as ‘other’. ‘Other’ was reserved for notes where clinicians were uncertain. As a result, these notes were dropped from the training set.

In addition to note labels, we aggregated note labels to generate patient labels. For binary diagnoses, the presence of a single glaucoma-diagnosing note was sufficient to label a patient as a ‘case’. For subtype diagnoses, we assigned the patient the label of their highest priority subtype present in their notes. This priority was defined as exfoliation spectrum, pigmentary spectrum, primary angle closure glaucoma, open angle glaucoma, glaucoma suspects, and ultimately no glaucoma.

### 4.3 Masked Pretraining

OphthaBERT was trained using continued masked pretraining on BioClinicalBERT in order to capture robust representations of ophthalmology notes. BioClinicalBERT was developed by further pretraining BioBERT, which was originally trained on a large corpus of tokens from PubMed, and later refined with data from the Medical Information Mart for Intensive Care III (MIMIC-III) dataset. In that phase, BioClinicalBERT learned from 2,083,108 de-identified notes related to hospital admissions between 2001 and 2012.

In masked pretraining, we randomly masked 15% of the tokens in each unlabeled note from the MEE corpus and tasked the model with predicting the missing tokens (Figure 6). To guard against overfitting, we used an 80/20 train/test split, set the initial learning rate to 5e-5, and trained for a maximum of 20 epochs with early stopping patience set at 2 epochs of no improvement. We trained with a batch size of 16 over 2 NVIDIA RTX6000 GPUs, utilized a linear scheduler with a warmup ratio of 5%, and applied a 10% weight decay.

**Figure 6:**
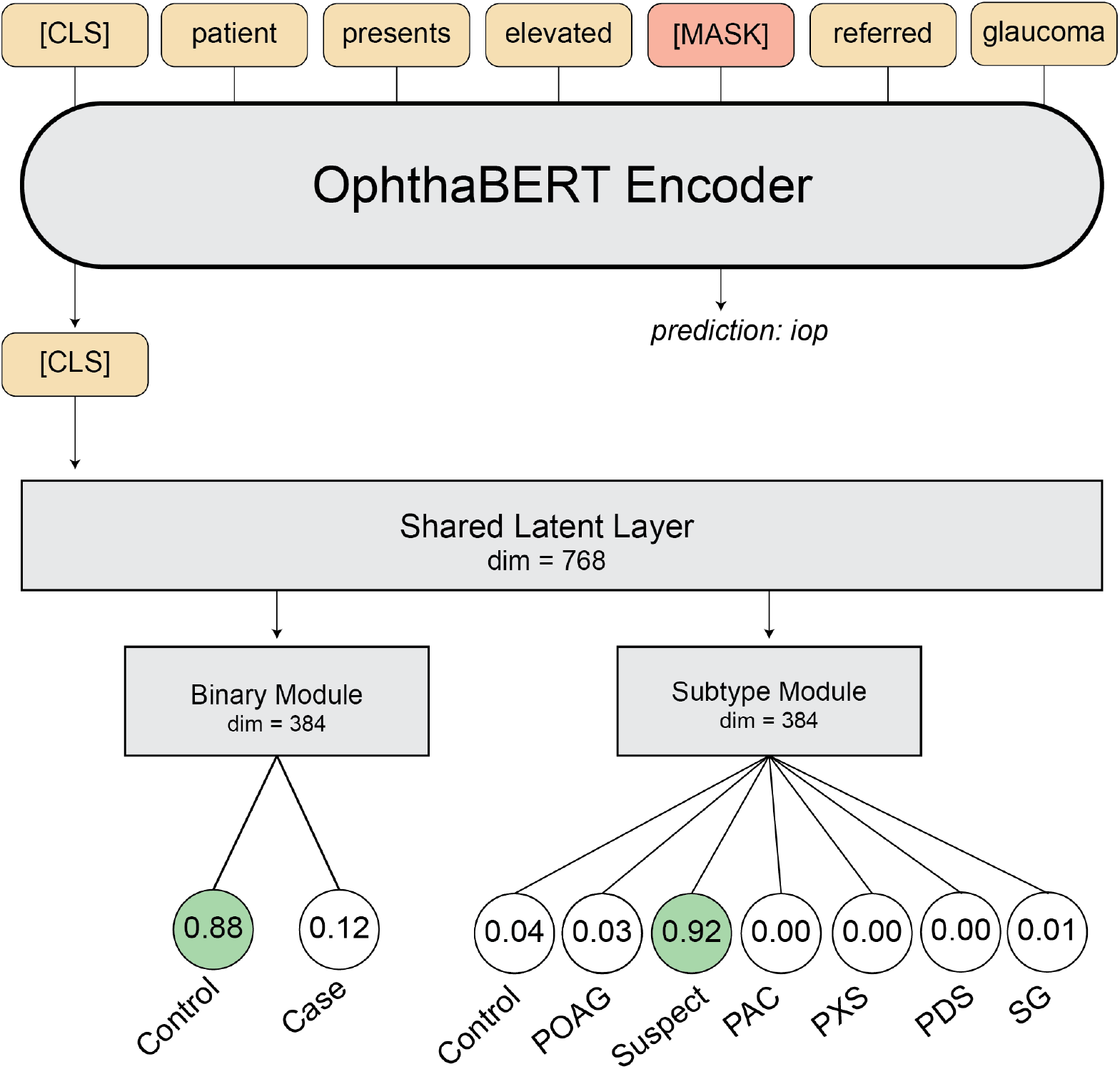
OphthaBERT Architecture. Architecture of the OphthaBERT model for masked pretraining and glaucoma diagnosis. Masked pretraining involves predicting missing tokens after 15% of the tokens are hidden from the model. The glaucoma model is built on top of the pretrained OphthaBERT model, with a binary classification module for binary diagnosis and a multiclass classification module for subtype identification. The model is trained using a joint loss function that averages binary cross-entropy loss and cross-entropy loss. The model learns shared representations for both tasks, allowing for efficient and effective classification. These shared representations are then fed into the classifiers.

### 4.4 Supervised Fine-tuning

We constructed the glaucoma variant of the pretrained OphthaBERT model by feeding the [CLS] token into a fully connected hidden layer of dimension 768, identical to the hidden layer size of BERT. A 20% dropout was applied to the shared hidden layer. This hidden layer was then connected to two separate pre-classification and classification heads: a binary classification head for glaucoma diagnosis and a 7-head classification head for subtype identification. Each classification head had a pre-classifier of dimension 384, which was fed into the respective classification head after a 10% dropout. A diagram of model architecture is available in Figure 6.

A ReLU activation function was used for all additional layers of the model, with a 20% dropout after the additional hidden layer. 10% dropout was applied after each preclassifier to the respective classification head. The loss function for supervised fine-tuning was defined as the average of the binary cross entropy loss from the binary head and the cross entropy loss from the subtype head:

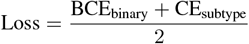

All parameters of OphthaBERT were fine-tuned through this joint loss function. We evaluated our model through 5-fold cross-validation, splitting by patient ID to ensure there is no leakage in the validation and testing set. We took 15% of training patients for validation and early stopping, keeping the testing set of each fold fully held out for evaluation after training completed. We augmented the training dataset using several techniques, including synonym replacement, abbreviation expansion, and sentence shuffling. Training was set at 50 epochs with an early stopping patience of 3 epochs. We set an initial learning rate of 3 × 10^−5^ and a batch size of 32. In training, we utilized a 10% weight decay and linear scheduler with a warmup ratio of 10%.

### 4.5 Model Evaluation and Interpretability

#### 4.5.1 Model Evaluation

We evaluated the performance of OphthaBERT using a variety of metrics, including accuracy, precision, recall, and F1 score. We also calculated the area under the receiver operating characteristic curve (ROC-AUC) and the area under the precision-recall curve (PR-AUC) to assess the model’s performance across different classification thresholds. In non-benchmarking results, we report the results of all testing sets in 5-fold cross-validation as described above.

We utilized McNemar’s test to evaluate the significance of the difference in binary glaucoma diagnosis performance for clinical notes before and after pretraining. To correct for multiple comparisons at different training data fractions, we utilize the Bonferroni correction to assess significance. As cross-validation is resource intensive for fine-tuning large models, we bootstrap sampled 95% confidence intervals of performance metrics for cross-model benchmarks on an 80/20 train-test split by patient ID.

#### 4.5.2 Benchmarking

We benchmarked OphthaBERT with several other models for glaucoma diagnosis. In order to make the benchmarking fair, we randomly sampled 20% of patients with labeled data to test the performance of each model. We utilized the same split for all models to ensure a fair benchmark. All notes were preprocessed using the standardized preprocessing as defined above, and patient labels were aggregated from note labels as defined above. For all models except OphthaBERT, two models were fine-tuned for binary and subtype classification respectively.

In random forest benchmarking, we represented our input features as TF-IDF bigram vectors with 5000 maximum features. Our random forest classifier was trained using warm-starting, allowing the number of trees to be incrementally increased based on validation performance.

We also fine-tuned MedAlpaca-7B, Gemma-1.1B-it, and Llama2-7B-it with low-rank adaptation (LoRA), setting the rank at 8 and the target modules as the query and value matrices. We utilized a warmup ratio of 5% and set the initial learning rate to 2e-4 with the AdamW optimizer. The model was allowed to converge fully for 10 epochs, with early stopping after 2000 steps of no improvement. We fine-tuned for causal language modeling with 16-bit floating point precision and a batch size of 1.

#### 4.5.3 External Validation

In addition to internal cross-validation and cross-model validation, we externally validated the binary classification module of OphthaBERT on a set of 70 notes from Wilmer Eye Institute and Johns Hopkins University (JHU). In order to ensure the model had access to sufficient information in the notes, we limited notes to overview notes, progress notes, or assessment and plan with overview notes. Assessment and plan notes alone were excluded. Instead, assessment and plan notes were concatenated with corresponding overview notes to maximize information available to the model.

No preprocessing was applied to the notes, and the notes were given binary labels and probabilities using the pretrained weights of OphthaBERT. A clinician at JHU assigned a label to each note: ‘Definitive Glaucoma’, ‘Possible Glaucoma’, or ‘No Glaucoma’. Due to the constraints of the grading team at JHU, subtype labels were not given to each note. In agreement with the binary labeling system in fine-tuning, ‘Possible Glaucoma’ and ‘No Glaucoma’ labels were merged into a single ‘Control’ label. We reported the performance of our model on this validation set for each category of notes when including and excluding ‘Possible Glaucoma’ notes.

#### 4.5.4 International Classification of Diseases (ICD) Codes Evaluation

We extracted the ICD codes for each patient in the labeled dataset. The priority defined for note labeling was used to assign a single ICD code to each patient. We then compared the performance of OphthaBERT with the ICD codes for each patient.

#### 4.5.5 Interpretability Analysis

Integrated Gradients was used to attribute specific tokens to class predictions in the fine-tuned variant of OphthaBERT for glaucoma identification [15]. We computed the attribution of each input token along a linear path from a baseline (zero) embedding to the true input embedding. The attribution for some token *x*_*i*_ is defined as:

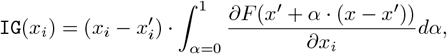

*F* (*x*) is defined as a function to represent the model’s predicted probability for a given class, and *x*^*′*^ represents the baseline input. We approximated this integral using a Riemann sum and ranked tokens based on their importance for predicting the defined class in a given note.

We evaluated interpretability by examining the attributions of the model for both binary and subtype classification on randomly sampled notes. We also averaged the attributions for all notes to assess the most important tokens for each class. Interpretability techniques were primarily used to assess overfitting.

## 5 Acknowledgements

This research was supported by NIH grant R01 EY036518.

## 6. Data Availability

In accordance with the policies of Massachusetts Eye and Ear, the data from this study cannot be publicly disclosed.

## 7. Code Availability

Private access to code and pretrained model weights will be made available upon request. All code and model weights will be made publicly available on GitHub and HuggingFace upon journal acceptance.

## 8. Ethics Statement

This study was approved by the Mass General Brigham IRB and Johns Hopkins Medicine IRB.

## 9. Competing Interests

N.Z. receives consulting fees from Sanofi.

## 10 Supplementary Material

## Notes

### Author Declarations

Ethical approval for this work was obtained from the Mass General Brigham Institutional Review Board (IRB) for Massachusetts Eye and Ear, and from the Johns Hopkins Medicine IRB for Johns Hopkins University.

